# Applying item response theory to psychometrically evaluate and shorten the Negative Acts Questionnaire-Revised

**DOI:** 10.1101/2024.09.21.24314135

**Authors:** Anna M. Dåderman, Petri J. Kajonius, Beata A. Basinska

## Abstract

Workplace bullying (WB) assessment often relies on the Negative Acts Questionnaire-Revised (NAQ-R). This study aimed to shorten and improve the NAQ-R using Item Response Theory (IRT) and address sex bias. IRT analysis from 867 Swedish employees (66% women) identified less-informative items. Based on this, a 13-item NAQ-R Short Form (NAQ-R-SF) was developed, demonstrating strong discrimination and validity. The new NAQ-R-SF showed a significant correlation with a primary WB measure (r = .57) and other relevant constructs, including individual factors like neuroticism and health quality, as well as work-related factors such as interpersonal conflicts and work performance. Sex bias was not found. IRT and validity evidence support the NAQ-R-SF as a robust tool for measuring WB, aligning with established WB constructs and individual differences.

## Introduction

This study focuses on the experience of workplace bullying (WB), specifically, the victimization aspect. WB, prevalent among colleagues or supervisors, manifests persistently and repeatedly toward an employee who lacks defense. One widely accepted definition of WB is provided by Einarsen et al. [1]:

> The term bullying refers to situations where an employee is persistently picked on or humiliated by leaders or fellow co-workers. A person is bullied or harassed when he or she feels repeatedly subjected to negative acts in the workplace, acts that the victim may find difficult to defend himself or herself against (p. 382-283).

Meta-analyses and reviews examining WB reveal its prevalence across industries and countries, exploring outcomes, mental health impact, and associated psychosocial factors [2]. They underline its frequency variation across workplaces and its diverse forms. WB is consistently linked to adverse mental health effects, including stress, anxiety, depression, and physical health issues [3–8], and to individual differences in personality traits [9,10]. These effects significantly impact the well-being of employees who experience WB, influencing their overall health quality and workplace functioning, such as interpersonal conflicts and work performance. Identifying patterns, predictors and risk factors—power imbalances, culture, and personality traits—guides intervention strategies.

The Negative Acts Questionnaire-Revised (NAQ-R) [11], a 22-item scale, remains prominent in WB measurement. Notelaers et al. [12] strongly advocate for shortening the NAQ-R and have delineated existing abbreviated measures derived from it. While these measures were developed using conventional Classical Test Theory (CTT) methods and expert consensus, none have utilized Item Response Theory (IRT) for abbreviation, despite its suitability for scale abbreviation. A condensed 9-item version (S-NAQ), as outlined by Notelaers et al. [12], has been distributed at two conferences to assess its applicability across various cultures. However, the process for selecting the items within the S-NAQ remains undisclosed. The selection process for the S-NAQ resulted in the exclusion of most items with the content related to work-related and physically intimidating bullying. Consequently, the short form (S-NAQ) primarily represents person-related bullying.

Expanding beyond the 9-item S-NAQ allows for a more detailed examination of WB. Additional items can explore specific nuances overlooked by the S-NAQ, providing richer data for intervention studies, ambulatory examination, or screening purposes. For instance, the S-NAQ fails to capture experiences such as having one’s opinions disregarded or encountering workplace humiliation. By evaluating all 22 items of the NAQ-R and selecting the most informative ones through item analysis, a comprehensive yet concise scale can be developed, ensuring reliable, effective and valid measurement of WB.

### Item response theory

IRT remains underutilized in WB research. IRT offers three primary advantages. Firstly, IRT models effectively utilize all available data, unlike confirmatory factor analysis (CFA) which relies solely on summary statistics [13]. Secondly, IRT models account for the ordinal nature of items and prioritize understanding the performance of each individual item [14].

Thirdly, reliability coefficients in classical test theory (e.g., Cronbach’s alpha) assume uniform standard error of measurement across the latent variable continuum. In contrast, IRT models integrate item characteristics and recognize that reliability of person scores may vary across different levels of the latent variable. Consequently, they can derive conditional reliability, which reflects reliability across the latent continuum [14].

Overall, IRT presents a data-driven and statistically robust method for shortening scales. In IRT, parameters *a* (item discrimination) and *b* (item location, also known as item threshold or item difficulty) are vital for evaluating individual item performance and overall evaluation effectiveness in measuring the intended latent trait. These parameters provide numerical insights into item behavior within the test, derived from responses of test takers.

Understanding these parameters aids in gauging item functionality and the information they convey about the latent trait being measured (e.g., WB). Utilizing IRT’s capacity to evaluate individual items, pinpoint redundant or problematic ones, and address issues like differential item functioning (DIF) while tailoring evaluations, IRT enables the development of shorter, more efficient, and precise scales compared to CTT [14]. Notably, the earlier formulated 9-item NAQ-R version, S-NAQ [12], lacked IRT methodology in its development, leaving uncertainties about the item selection process. In our study, we apply IRT to evaluate the 22-item NAQ-R scale, selecting the most informative items to create a shortened version, while also evaluating the existing 9-item scale with the goal of enhancing its quality. IRT application in shortening the NAQ-R or identifying possible sex-related biases in response patterns is notably scarce.

### Unexplored differential item functioning in NAQ-R

The evidence regarding sex-related differences in victimization rates presents a complex picture. While certain studies emphasize a higher incidence of victimization among females, conflicting findings exist. For instance, while some studies indicate a prevalence of female victims over males [15], Notalaers et al. [16] pointed out that among 15 studies, four reported more female victims. Consequently, these comparisons might be misleading as it is unclear whether the divergence in NAQ-R outcomes represents an authentic distinction or stems from factors like different interpretations of survey items between men and women. It may be evaluated by DIF in a scale. Significantly, research on sex biases in NAQ-R responses is scarce. However, Sischka et al. [17] found no sex-related measurement differences in another WB measure. Hence, we didn’t expect sex-related disparities in NAQ-R interpretation. We assert that conclusive evidence on this matter is still lacking. DIF may stem from varied interpretations of survey items and diverse bullying experiences across sexes. Organizational members might display unique negative behaviors towards men and women due to gender stereotypes. For instance, women’s opinions may be dismissed more often, while men may face more practical jokes. In summary, crucial gaps in current research on NAQ-R include utilizing advanced methodologies such as IRT for refining measurement approaches, and addressing potential sex-related bias.

### Current study

Building upon the overview provided earlier, this study aims to address two critical gaps in the existing literature. Firstly, our study focuses on shortening the still widely-used 22-item NAQ-R. We apply methodologies explicitly designed to identify the most informative items and evaluate the item quality chosen for the creation of the relatively recently published 9-item S-NAQ [12]. Secondly, it aims to discern whether in some studies observed sex-related differences in WB are authentic or stem from psychometric variations in certain NAQ-R items between men and women.

The primary objectives of this study were to apply IRT to psychometrically analyze the items of the NAQ-R and S-NAQ, assess the overall scale properties, and create an abbreviated version of the NAQ-R. Additionally, we aimed to investigate potential DIF across sexes. We also sought to evaluate the comparability of the 22-item NAQ-R, the 9-item S-NAQ, and the newly developed abbreviated version, the NAQ-R-SF, in terms of key psychometric properties such as item parameters, reliability, and concurrent validity. Additionally, we assessed the convergent and divergent validity of the NAQ-R-SF by examining its correlations with constructs highlighted in WB meta-analyses, including individual differences in health quality, personality traits, and workplace functioning (e.g., interpersonal conflicts and job performance). Our study successfully met these objectives.

## Materials and methods

### Participants and procedure

The study included employees from various organizations in Sweden. The majority (60%) worked in social services, healthcare, and welfare, while the remaining participants were employed in diverse professions such as technical roles, restaurant management, office work, teaching, and security. Participants did not receive any compensation.

Data were collected initially through paper-based methods from January 1^st^, 2015 (*n* = 204) and electronically from January 1^st^ 2015 to December 31^st^ 2019 via social media (*n* = 663) using a snowball sampling technique. Written individual consent was collected at the start of the survey. The online and pencil-and-paper versions of the questionnaire, NAQ-R, showed no differences in content, delivery, functionality, or user experience. A preliminary evaluation of the psychometric equivalence between the two versions of the 22-item NAQ-R was conducted. However, response bias and test-taking strategies were not measured. It is possible to consider both versions to be approximately psychometrically equivalent. The Cronbach’s alpha for the pencil-and-paper version was .90, and for the online version, it was .93. Both versions showed comparable correlation values between NAQ-R and a single-item measure of WB (*r* = .68 vs. .61, *z* = 1.49, *p* = .068).

Demographics were obtained across these instances, forming the basis for the overall sample description. Participants, aged 17–75 (*M* = 39.0, *SD* = 11.4), constituted 66% women. Education levels varied: 34% completed upper secondary education, 22% had < 3 years of higher education, and 44% had ≥ 3 years. The majority (63%) were married or cohabiting, with professional experience spanning from 0.1 to 41 years (*M* = 7.1, *SD* = 7.0). They typically worked in groups ranging from 1 to 50 members (*M* = 16, *SD* = 10). Most (74.5%) worked full-time.

### Ethical statement

The study was conducted in accordance with the Swedish Ethical Review Act (SFS 2003:460). Prior to commencing data sampling in 2015, this study underwent consultation with a scientific secretary at the former Regional Ethical Board, now known as the Swedish Ethical Review Authority. Formal approval by the Ethical Review Authority was not required for this study, as it focuses solely on psychometric analysis of an anonymous questionnaire, without involving experiments or sensitive data usage. All protocols for methods and analyses were in line with Lund University’s internal ethical guidelines. Data collection did not involve manipulation or deception tactics, and was conducted voluntarily. It involved anonymous standardized questionnaires, ensuring participant confidentiality and adherence to ethical standards. Written consent was obtained following the Declaration of Helsinki.

### Measures of workplace bullying

#### Negative Act Questionnaire-Revised (NAQ-R)

To examine WB experiences over the past six months, this study utilized the NAQ-R [11], which was translated from Norwegian to Swedish, adapted, and published online by Dåderman and Ragnestål-Impola [18]. The NAQ-R consists of two distinct parts: the first includes 22 items, while the second comprises a single-item measure of WB (Item 23). The second part provides a definition of WB and asks about the respondent’s subjective experience of being bullied.

The first part involves objectively worded items probing experiences of negative acts at work across three different situation-related negative forms of behaviors: work-related (seven items, e.g. “Having your opinions ignored”), person-related (12 items, e.g. “Being ignored or facing a hostile reaction when you approach”), and physically intimidating (three items, e.g., “Intimidating behaviors such as finger pointing, invasion of personal space, showing, blocking your way”). The items are framed in behavioral language without explicitly mentioning the term “bullying.” The Swedish version by Dåderman and Ragnestål-Impola deviates slightly from the original Norwegian version by Einarsen et al. [11]. For instance, the NAQ-R’s response format uses a Likert-like frequency-based scale (1 = “never”, 2 = “now and then”, 3 = “monthly”, 4 = “weekly”, and 5 = “daily”). Caponecchia and Costa criticized this format for its inconsistent intervals and the ambiguity of the “now and then” option. In the adapted Swedish version published by Dåderman and Ragnestål-Impola in 2019, the response option “now and then” was replaced with “sometimes” to alleviate interpretational ambiguity, addressing the critique that “now and then” could be misconstrued due to its placement between “never” and “monthly.” This change holds significance as it transitions the response format into a Likert-like ordered scale, rendering it more suitable for analysis not only through IRT but also via metric models like factor analyses [20]. Furthermore, in the adaptation by Dåderman and Ragnestål-Impola, Item 6 (“Exclusion from the social community”) omitted the idiom “sent to Coventry” as it was criticized by Fevre at al. [21] for its lack of universality. Fevre et al. also criticized Items 18 and 20 for including the term “excessive,” which can be widely interpreted. In the adaptation by Dåderman and Ragnestål-Impola, this term was revised to “unreasonable”.

Research on the NAQ-R’s factor structure reveals varied outcomes, with a prevalent single-factor model indicating WB. Some suggest two or three factors [11], representing distinct forms of WB, but these factors have very high latent intercorrelations (person-related with work-related *r* = .96 and with physically intimidating bullying *r* = .89). Replication across diverse samples has been inconsistent. Most studies treat WB as a unidimensional construct [22,23] to capture the broader WB experience, a perspective also adopted in the current study.

#### Single-item measure of workplace bullying

The second part of the NAQ-R, titled “About bullying,” consists solely of a single-item measure of WB (Item 23), which aligns with Einarsen et al.’s [11] definition of WB. This single-item measure asks respondents, “Have you been bullied at your workplace?” The provided definition of bullying encompasses repeated exposure to unpleasant, degrading, or peculiar treatment at work, lasting for a certain period and causing difficulties in self-defense. Response options range from 1 (“no”) to 5 (“yes, daily”). This single-item measure of WB is designed to assess the respondent’s personal experience of WB. In this study, it was used to assess concurrent validity of the new 13-item NAQ-R-SF developed in the current research.

#### Short Negative Act Questionnaire (S-NAQ)

The 9-item S-NAQ was derived from the 22-item NAQ-R through CTT, discussions at two International Association of Workplace Harassment and Bullying conferences, and validation using latent class analysis. Unlike IRT, which is specifically designed for shortening assessment tools, the S-NAQ was not reduced using such techniques. Our study applied IRT to evaluate the S-NAQ.

### Measures used to confirm validity of the NAQ-R-SF

We included measures to confirm the convergent and divergent validity of the new NAQ-R-SF. These measures are short versions of Swedish adaptations of key constructs identified in WB meta-analytic research. These constructs encompass individual differences in experiencing WB, such as health quality and personality traits, as well as variations in workplace functioning, including interpersonal conflicts and work performance.

#### EuroQol Five-Dimension Questionnaire (EQ-5D-3L)

Given the extensive empirical research highlighting relationships between WB and employee health-related well-being [4,6–8], we included the EuroQol Five-Dimension Questionnaire (EQ-5D-3L) [24], a generic health-related quality of life instrument. Over a third of the participants (37%; *n* = 324) completed its officially translated Swedish version (EQ-5D-3L | EuroQol).

The EQ-5D-3L has two parts. The first is a 5-item questionnaire assessing health-related quality of life across five dimensions: mobility, self-care, usual activities, pain/discomfort, and anxiety/depression, with responses from “no difficulties” to “extreme difficulties.” Scores form a five-digit health profile, convertible into a utility index using Swedish data [25]. The second part, the EQ-VAS, is a vertical scale where respondents rate their overall health quality from 0 (worst) to 100 (best). Both parts were used in this study.

#### Mini International Personality Item Pool-6 Inventory (Mini IPIP6)

The individual disposition hypothesis suggests that certain personality traits, such as neuroticism, may predispose an employee to experience WB [10]. Neuroticism is both a well-established antecedent and consequence of WB. A meta-analysis by Nielsen et al. [9] on workplace harassment, a broader concept than WB, found harassment positively associated with neuroticism (*r* = .25) and negatively associated with extraversion (*r* = −.10), agreeableness (*r* = −.17), and conscientiousness (*r* = −.10), with no significant relationship to openness (*r* = .04). However, more recent research [18,23] indicates that openness is also negatively correlated with WB. Openness may serve as a moderator in the relationships between WB and health-related quality of life [26]. In this study, most participants (88%, *n* = 767) completed the Mini-IPIP6 [27].

The Mini-IPIP6 is a 24-item personality assessment tool, evaluating six traits: extraversion, agreeableness, conscientiousness, neuroticism, openness, and honesty-humility, with 4 items dedicated to each trait. It uses responses ranging from 1 = “strongly disagree” to 7 = “strongly agree”. The Swedish version (translated and adapted by Backström, Dåderman, Grankvist, Kajonius, and Lundin) is published online [18]. Extraversion involves energy, sociability, talkativeness, and assertiveness. Agreeableness includes kindness, helpfulness, and cooperation. Conscientiousness covers organization, reliability, and goal-orientation. Neuroticism indicates worry and anxiety. Openness reflects imagination and curiosity. Honesty-humility represents fairness and genuine behavior, even when exploitation is possible [28]. In this study, Cronbach’s alphas (α) and mean inter-item correlations (*M*_iic_) were: extraversion (.79/.36), agreeableness (.75/.43), conscientiousness (.76/.44), neuroticism (.69/.26), openness (.63/.32), and honesty-humility (.67/.38).

#### Interpersonal Conflict at Work Scale (ICAWS)

WB can sometimes be referred to as coworker conflict. Spector and Jex [29] describe workplace interpersonal conflicts as ranging from minor disagreements to physical abuse, distinguishing between open conflicts (e.g., rudeness) and covert conflicts (e.g., rumor-spreading). Their findings indicate that such conflicts can disrupt workflow, hinder task cooperation, and lead to role conflicts, intentions to resign, as well as anxiety and depression. Employees who experience WB often report anxiety, depression, intentions to leave the workplace, and role conflicts. In this study, over a quarter of participants (27%; *n* = 231) completed the ICAWS [29] (α = .80, *M*_iic_ = .49).

ICAWS is a 4-item measure of the frequency of conflict behaviors at work over the past month on a 5-point Likert-type scale (1 = “never”, 5 = “very often”). The Swedish version was translated by Granqvist and back-translated by Lundin. Its validity was confirmed through strong correlations with work-family conflict in workplace settings [30].

#### Individual Work Performance Questionnaire (IWPQ)

Strong meta-analytic evidence shows that employees who experience WB report high levels of mental distress and lower well-being [8], and they also score low on their work performance [7,30]. Research by Devonish [31] revealed that WB was negatively associated with task performance (*r* = −.30) and interpersonal organizational citizenship behavior (*r* = −.29) and positively associated with interpersonal counterproductive work behavior (CWB; *r* = .43). About a quarter of the participants (24.5%; *n* = 212) completed the IWPQ [32], measuring task performance (α = .68, *M*_iic_ = .30), contextual performance or organizational citizenship behavior (α = .83, *M*_iic_ = .39), and CWB (α = .77, *M*_iic_ = .39).

IWPQ is an 18-item measure of individual work performance using responses ranging 1– 5, from “seldom” to “always” for task and contextual performance, and from “never” to “often” for CWB. All items have a recall period of 3 months. The Swedish version of the IWPQ has been published and validated [33,34]. Task performance involves meeting job expectations in quantity, quality, essential skills, and professional knowledge, including planning, problem-solving, accuracy, knowledge maintenance, goal setting, and timely goal achievement. Contextual performance extends beyond duties, involving extra tasks, project initiation, collaboration, offering advice, and enthusiasm. Conversely, CWB harm the organization, including complaints, negativity, off-task behavior, presenteeism, intentional mistakes, misuse of privileges, and exaggerating challenges.

### Preliminary tests evaluating IRT assumptions

The IRT analysis utilized 2PLM IRT for Patient-Reported Outcomes (IRTPRO), and in accordance with the NAQ-R’s fem Likert-like response categories (1-5), a graded response model (GRM) [35] was selected. Prior to IRT, three key assumptions were scrutinized: approximately unidimensionality, monotonicity, and item independence.

The concept of approximate unidimensionality suggests that a test or set of items measures a single underlying latent trait, although strict adherence to this assumption is not always necessary. Reckase [36] demonstrated that one dominant factor significantly influencing item responses is often adequate for analysis. In simpler terms, the test should evaluate one central construct rather than multiple unrelated ones. This is evaluated by using exploratory factor analysis (EFA). Commonly used indicators supporting approximate unidimensionality include: (a) the first factor explaining at least 20% of the variance [36]; or (b) a ratio greater than 3 between the eigenvalues of the first and second factors [37].

IRTPRO does not feature a specific test for evaluating monotonicity directly. However, potential violations can be indirectly evaluated by examining item response functions. In our study, we applied the Mokken scale analysis [38] in R package mokken (version 3.1.0), which provides a detailed breakdown of individual items and their contribution to the scale’s measurement quality, specifically in terms of the monotonicity assumption in IRT. For example, the Mokken scalability coefficient (H-coefficient) gauges the extent to which each item adheres to the monotonicity principle. Higher values of this coefficient indicate stronger evidence supporting monotonicity for that particular item. According to Van der Ark [38], a coefficient greater than .30 is indicative of satisfactory adherence to monotonicity. Understanding the contribution of each item to the overall functioning of the scale is what renders Mokken analysis invaluable for scale development and refinement.

IRTPRO software offers functionalities to evaluate local dependence (LD) [39]. LD can occur, for example, when the wording of two or more items is similar or uses synonyms, making it difficult for participants to distinguish between the items. As a result, they may select the same response category for all items. The evaluation of LD involves examining marginal fit (X^2^) and standardized LD X^2^ statistics, also known as the Chen and Thissen LD X² statistics. This statistic quantifies the level of dependence between two items by computing the squared difference between their observed and expected covariances, then dividing by the expected variance of the covariances assuming independence. Values that are high (e.g., exceeding 10) indicate a significant level of dependence, indicating that the items may be measuring distinct constructs or inappropriately influencing each other. While standardized LD X² aids in identifying potential dependencies, it is essential to complement it with other methods and expert judgment to draw informed conclusions. This may entail scrutinizing the content of the items. We examined both the content and factor loadings of pairs of items exhibiting LD to identify strong candidates for removal from the NAQ-R.

### Data management, analyses and modelling

Prior to aggregation, data were meticulously cleaned. Some IRT models can partially handle missing data by estimating item parameter levels from observed data, accommodating missing responses for individuals or items. We have chosen to pursue our objectives with a complete dataset for thoroughness, and applied a straightforward approach for handling missing data (< 1%): mean item imputation, rounded to the nearest whole values.

When a scale has fewer than eight response options, Cronbach’s alpha may be inappropriate for measuring reliability. Therefore, we calculated the mean inter-item correlation, ideally between .20 and .40. Additionally, to compare the correlation coefficients, we used the [Online-Calculator for testing correlations: Psychometrica](https://www.psychometrica.de/correlation.html#Testing%20Linear%20Independence). These correlations were derived from the same sample, leveraging this dependence to increase the power of the significance test.

Like other measures assessing traits such as psychopathy, psychiatric disorders, and socially negative behaviors prevalent in only a small percentage of the general population, we anticipated skewness in the NAQ-R. Given our sizable sample size, we did not anticipate skewness to compromise our analyses. Based on the central limit theorem, it is observed that when large samples are drawn from skewed populations, the resulting means tend to conform to a normal distribution [20]. However, compromising and acknowledging lower power of nonparametric tests we opted for Spearman’s coefficient over Pearson’s coefficient when evaluating the concurrent validity.

Initially, we estimated an IRT model-data fit at both item and model levels using 22 items with IRTPRO. Then, we estimated IRT model-data fit for both short versions of the NAQ-R. We evaluated the absolute fit of the model to each item, using a generalization of Orlando and Thissen’s [40] S-χ^2^ item-fit statistics for polytomous data. Item-fit statistics were evaluated at 1% significance level, as recommended by Stone and Zhang [41]. Model-data fit was evaluated by χ^2Loglikelihood^, limited information goodness-of-fit statistic correcting for sparse information in one and two-way marginal tables (*M*^2^) [42] and its associated *p* value, root mean square error of approximation (RMSEA) [43], and error prediction estimates via Akaike information criterion (AIC) [44] and Bayesian information criterion (BIC) [45].

Toland [46] detailed the steps for conducting IRT analyses and explained the interpretation of S-χ2 item-fit statistics and *M*^2^ limited information goodness-of-fit statistic as provided in IRTPRO. Briefly, like other goodness-of-fit statistics, *M*^2^ assumes perfect model-data fit in the population. Due to its sensitivity to minor model-data misfits, a nonsignificant *p*-value is not expected. Smaller *M*^2^ values indicate better fit. RMSEA is defined similarly to its use in CTT [47].

Subsequently, discrimination (*a*) and location (*b*) item parameters were examined, guiding the selection of the most informative items to compose the new NAQ-R-SF.

DIF statistics using Wald tests [48] identifies non-invariance (*p* < .05), anchoring invariant items while evaluating non-invariant ones. These analyses were conducted for the 22-item NAQ-R and for both 9-item S-NAQ and 13-item NAQ-R-SF measures. We applied the Bonferroni correction to adjust for multiple testing to control Type I errors in our results.

To further validate our results and enable comparison with other researchers—specifically those who have treated the NAQ-R as a continuous unidimensional total scale score—we applied three single-factor CFA models. These models were applied to the 22-item NAQ-R, the 9-item S-NAQ, and the 13-item NAQ-R-SF. These analyses, conducted using AMOS software with the maximum likelihood estimation method, aimed to verify the approximate similarity between the three versions of the NAQ-R. We allowed to correlate residuals based on modification indices and substantive analysis of the items. All versions represent the same underlying data and use the same response format.

To assess the concurrent validity, the NAQ-R-SF was correlated with the single-item WB measure, NAQ-R, and S-NAQ. To examine the convergent and divergent validity of the NAQ-R-SF we evaluated key constructs identified in WB meta-analytic research. Individual differences in experiencing WB, such as health quality and personality traits, were examined by correlating the NAQ-R-SF with the EQ-5D-3L, and the six personality traits from the MiniIPIP-6. Variations in workplace functioning, such as interpersonal conflicts and work performance, were examined by correlating the NAQ-R-SF with the ICAWS, and the three types of individual work performance from the IWPQ.

### Item response theory model

The GRM [35] was applied, encompassing discrimination (*a*) and location (*b*) parameters within the IRT analysis. Item quality was evaluated through the discrimination and location parameters estimated for each item. Item Characteristic Curves (ICCs) depicted the connection between an individual’s position on the latent trait (in this case, WB) and their likelihood of responding to an item designed for WB. Furthermore, item quality was evaluated based on the ICCs, which could be transformed into item information—a higher information value *a* indicating superior item quality. Aggregating information across all items yielded the test information, serving as an index for evaluating test precision. IRT offers a unique advantage in that it allows for the computation of two types of reliability: Conditional reliability, which accounts for potential variations in reliability across different levels of the latent variable, and marginal reliability coefficients, akin to overall reliability measures found in CTT frameworks such as Cronbach’s alpha.

### Construction and psychometric evaluation of the NAQ-R-SF

The new 13-item NAQ-R-SF was developed by removing from the NAQ-R items exhibiting relatively inferior item parameters compared to others. Evaluation of the psychometric properties of the short form (NAQ-R-SF) encompassed various analyses: unidimensionality testing, model-data fit analysis of the IRT model, evaluation of local independence, estimation of item parameters *a* and *b* and factor loadings λ, internal consistency and marginal reliability examination, evaluation of test information, and validity.

We exclusively provide illustrative data for the NAQ-R, as both abbreviated versions are derived from this comprehensive measure, incorporating subsets of its items. Item Information Functions (IIF; dashed lines) indicate how much empirical information (precision) each item contributes to the entire measure and where along the continuum this information is provided. The Test Information Function (TIF) represents the sum of the areas under each IIF, reflecting both the unique amount of information each item provides and the total number of items.

## Results

### Preliminary analyses: evaluating IRT assumptions

We first evaluated the assumption of approximate unidimensionality. The first eigenvalue (9.4) substantially exceeded the second (1.7), with a ratio (> 4) favoring unidimensionality. Moreover, the first factor explained 43% of total variance, significantly more than the second (8%), further supporting approximate unidimensionality.

The Mokken analysis, used to evaluate monotonicity, revealed that only Item 22 had a low scalability H-coefficient of 0.25, with no other identified violations. Item 3 (“Being ordered to do work below your level of competence”) displayed a H-coefficient of 0.32, exhibiting the highest values among all items for various metrics including the total number of active pairs (#ac = 112), total number of violations (#vi = 8), average number of violations per active pair (#vi#ac = 0.07), maximum violation (maxvi = 0.07), sum of all violations (sum = 0.41), average violation per active pair (sum/#ac = 0.0037), and maximum test statistics (zmax = 1.99). Given these findings, especially the significant violation observed, Item 3 appears to be a strong candidate for removal from the item set of NAQ-R due to concerns regarding monotonicity (Fig. 1).

**Fig 1.**
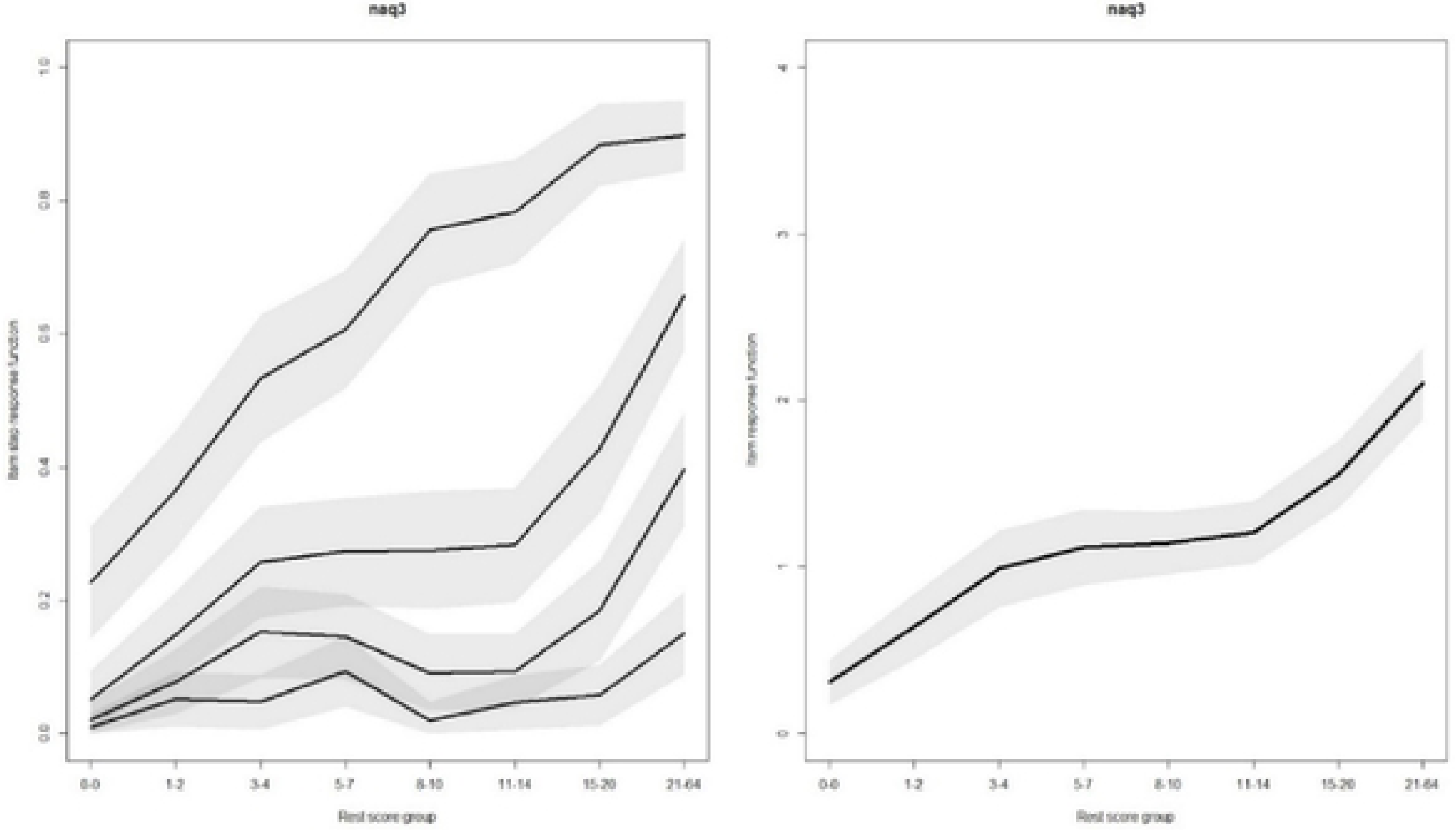
Visualizing a Violated Response Step Function in Item 3. Item 3 of the NAQ-R: “Being ordered to do work below your level of competence”. The horizontal axis depicts the latent trait (in this case, workplace bullying) of the 867 respondents of the NAQ-R, ranging from low to high values. The vertical axis represents the probability of endorsing each step of Item 3, ranging from 0 (never endorsing) to 1 (always endorsing). Each line corresponds to a step within the polytomous Item 3. The shaded area indicates the confidence interval around the estimated functions. While the lines typically show increasing functions of the latent trait, Item 3 notably violates the assumption of monotonicity.

Finally, we evaluated the assumption of item independence. Several LD X^2^ statistics surpassed 10, suggesting potential LD. Specifically, the following item pairs displayed LD: Item 3 (see above) and Item 17 (“Having allegations made against you”); Item 22 (“Threats of violence or physical abuse”) and Item 9 (“Intimidating behaviors like finger-pointing, invasion of personal space”); and Item 21 (“Exposure to an unmanageable workload”) and Item 16 (“Tasks with unreasonable deadlines”). Notably, the latter pair demonstrated the highest LD X^2^ value of 44.4, while the others were under 15. After analyzing content of the pairs of items exhibiting potential LD, we decided that items showing lowest λ in each pair would be good candidates to be removed from the item set of the NAQ-R. The items were: Item 3, 22 and 16.

### IRT analyses

Table 1 presents IRT model-data fit results, including item parameters (*a* and *b*), factor loadings, and fit statistics for the 22-item NAQ-R, 9-item S-NAQ, and 13-item NAQ-R-SF derived in this study. Variations in discrimination, location, and reliability among NAQ-R items were identified through IRT analyses. Table 1 shows that the S-χ2 item-fit statistics for the NAQ-R indicated a satisfactory fit on the item level, with only 4 out of 22 items not well represented by the estimated item parameters. Items 3 and 4 are no longer part of the short versions, S-NAQ and NAQ-R-SF. The remaining items with poor S-χ2 item-fit statistics in the NAQ-R showed satisfactory fit in these short versions. Item-level fit results indicated that items in these short versions had adequate fit, except for Items 2 and 5.

**Table 1.**
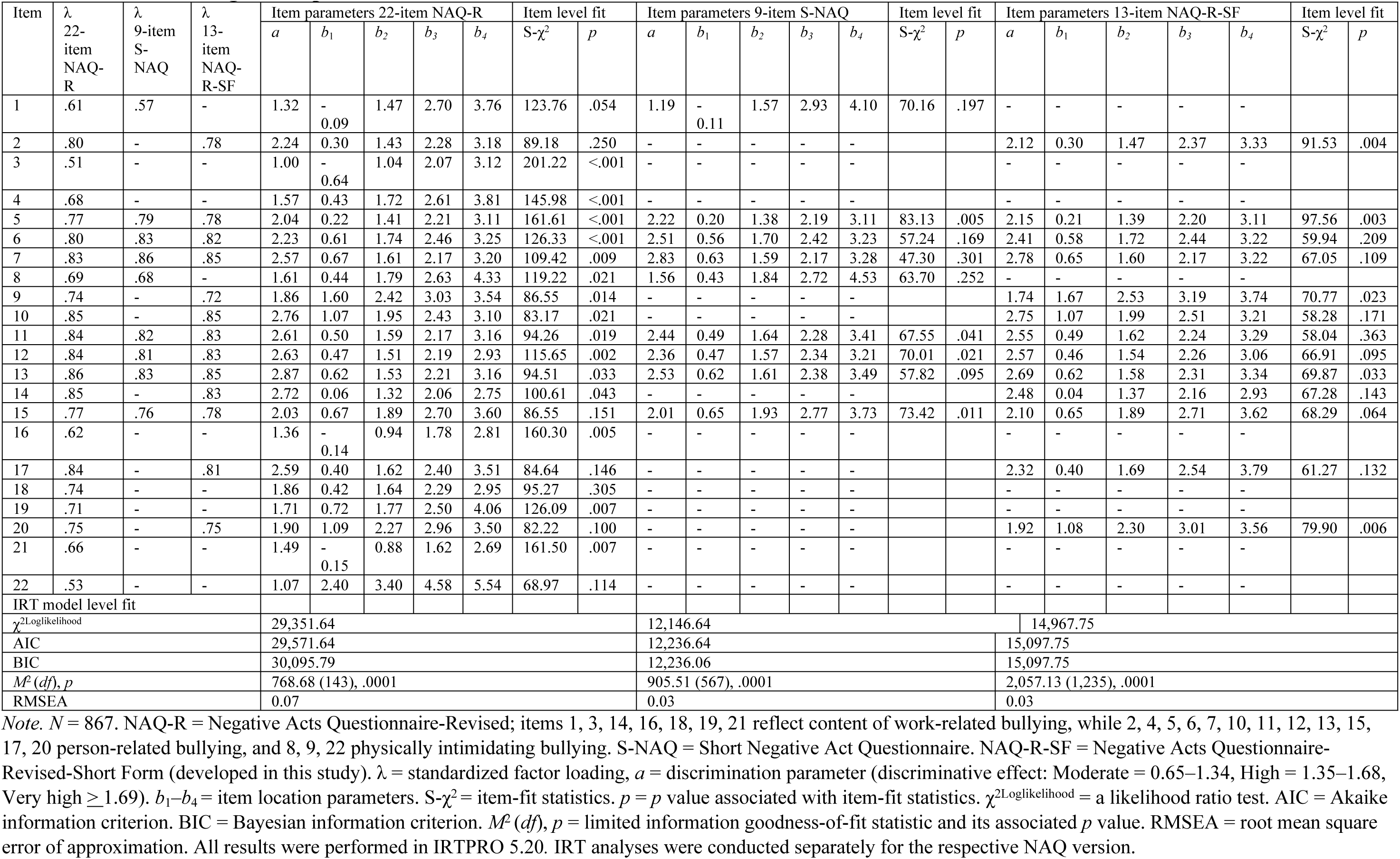
Factor loadings, item parameters, and model-data fit statistics of the NAQ-R, S-NAQ, and the NAQ-R-SF.

As expected, most model level fit statistics did not fit the data exactly, because they assume perfect model-data fit in the population. However, the RMSEA indicated similar and adequate model-fit for the three versions of the NAQ-R; it was better for the short versions.

The results shown in Table 1 indicate that the 13-item NAQ-R-SF comprises items with exceptionally high discriminatory power, representing the most informative items regarding parameters *a* and *b*.

In our comparative analysis utilizing IRT, we evaluated the 9-item version S-NAQ. Our investigation reveals that two of the nine items, specifically Item 1 (related to work-related bullying) and Item 8 (related to physically intimidating bullying), exhibit inefficacy in comparison to the other items in this version, as well as in contrast to both the 22-item NAQ-R and the 13-item NAQ-R-SF. These items displayed lower discriminative effect (*a* =1.19 and 1.56, respectively), and lower factor loadings (λ = .57 and .68, respectively).

While *a* and *b* parameters provide valuable insights, they should not be the sole criteria for evaluating item quality. Item 19 (“Pressure not to claim something to which you are entitled by right, e.g., sick leave, holiday entitlement, travel expenses”) demonstrated favorable *a* and *b* parameters; nevertheless, we conducted a content evaluation of all items. We opted to exclude Item 19 due to robust legal protections against such practices in Sweden.

Table 1 indicates consistent quantity and ratio of items addressing work-related and physically intimidating bullying in both abbreviated versions. Each content type is represented by one item, chosen more aptly through IRT compared to traditional methods. The condensation resulted in a 64% reduction in work-related bullying items and an 86% reduction in physically intimidating bullying compared to the 22-item NAQ-R.

IRT results for the 22-item NAQ-R are visually depicted in Figs 2 and 3. We analyzed the item properties, including the amount of psychometric information (precision) available for each NAQ-R item or subset of items (Fig 2), and for the entire measure (Fig 3).

**Fig 2.**
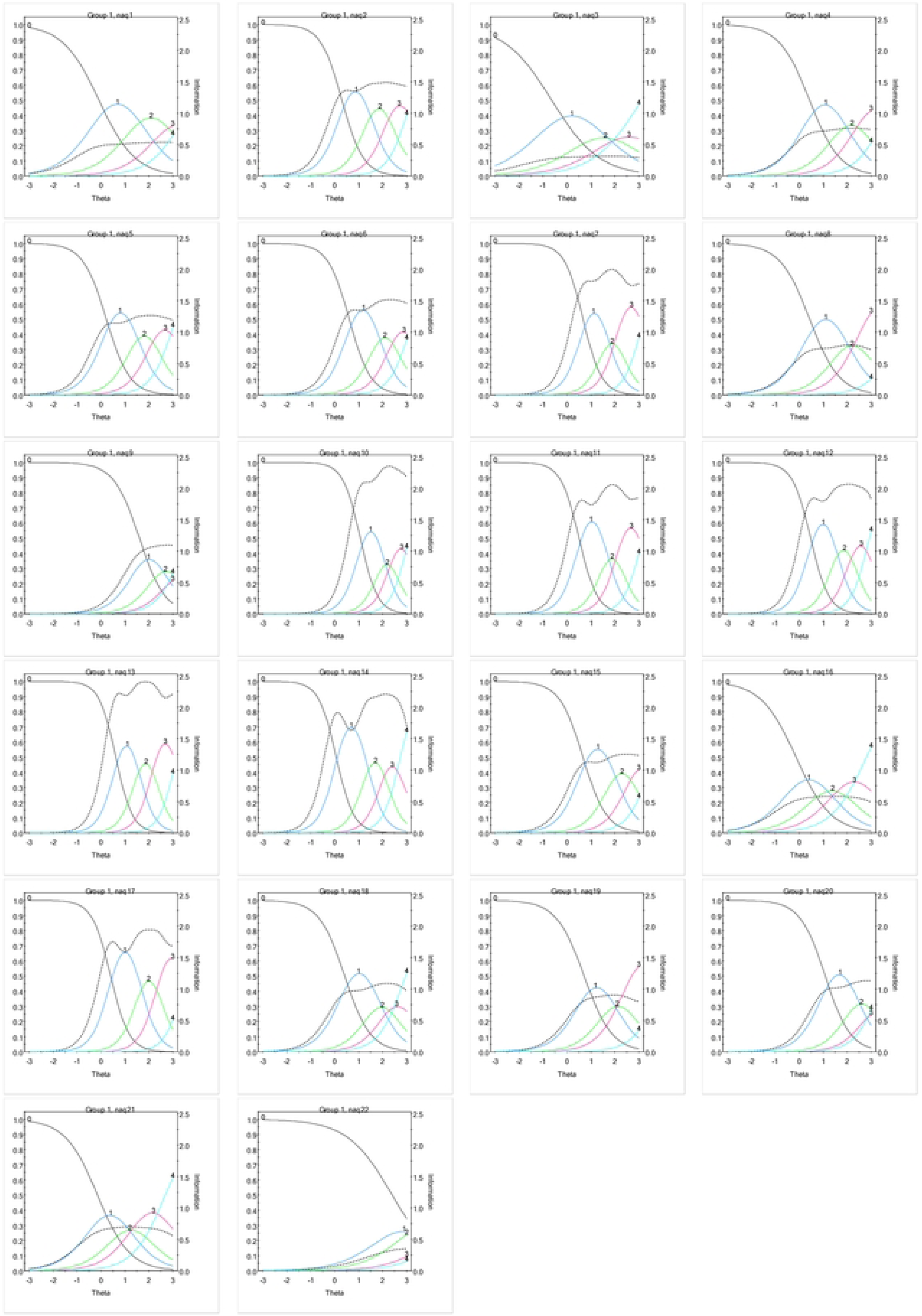
Item characteristics curves (ICC; colored lines) combined with item information functions (IIF; dashed lines) for each of the 22-items comprising NAQ-R (*N* = 867). Labeling the sample as “Group 1” indicates that it has not been visualized with regard to subgroups, such as men and women. Each figure contains colored and dashed lines corresponding to different items in the NAQ-R. These lines, representing Item Characteristic Curves (ICCs) in color and Item Information Functions (IIFs) in dashed lines, offer graphical representations used to analyze item behavior in IRT models. Colored lines indicate how the probability of the respective response changes across the WB range, while dashed lines illustrate the amount of information the item contributes to estimating the WB level of all responders with varying WB levels. By examining both ICCs and IIFs simultaneously, valuable insights can be gained into each item’s characteristics, including item location (also known as “difficulty” or “threshold”), discrimination, and information. (See Fig. 1 for the description of horizontal and vertical axes.)

**Fig 3.**
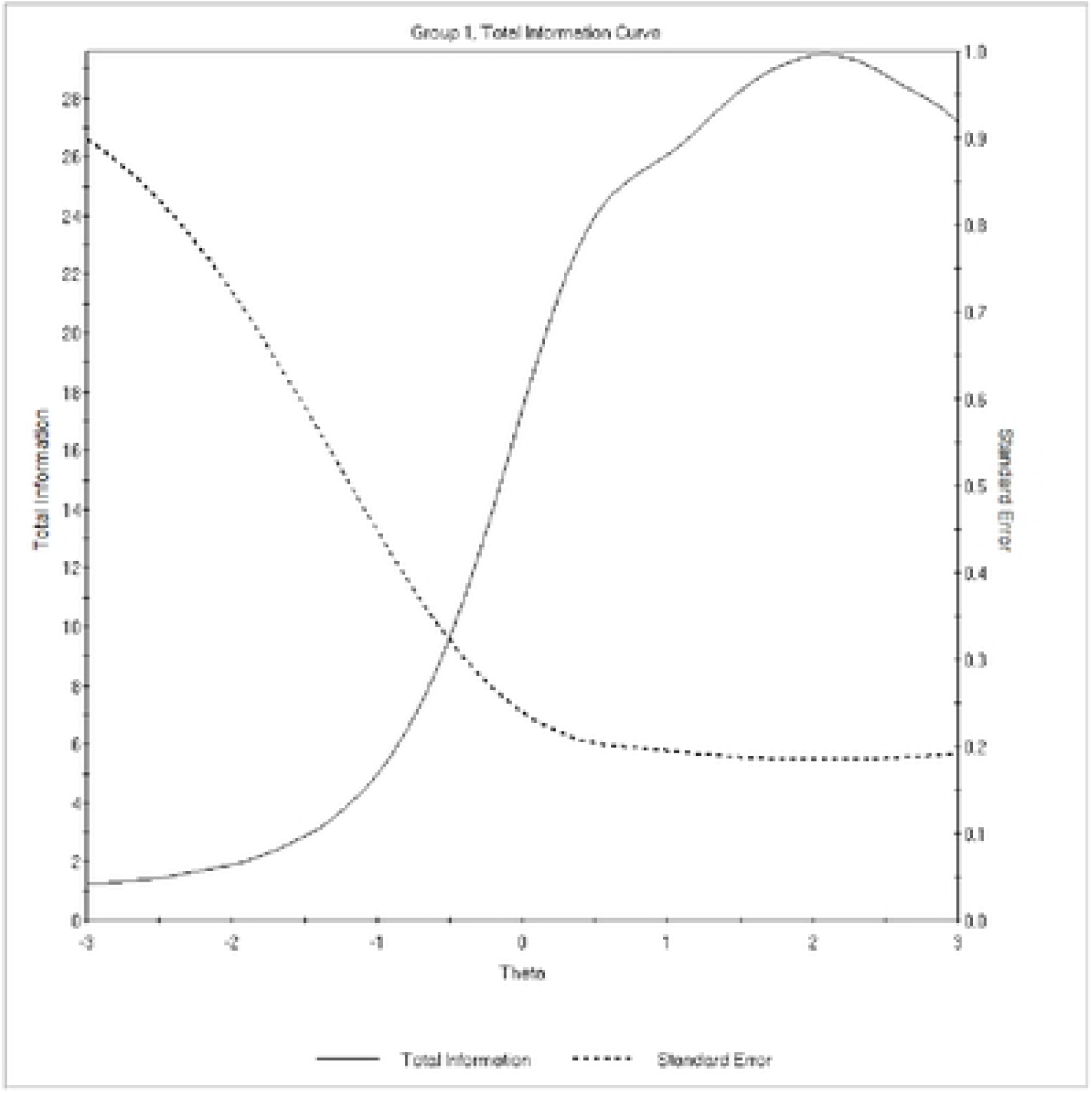
Test information function (TIF) of the Workplace Bullying by 22-item NAQ-R under the graded response model (*N* = 867) showing marginal reliability. The horizontal axis illustrates the latent trait θ of workplace bullying (WB), while the vertical axis represents the amount of information and the standard error provided by the NAQ-R across various levels of WB. Ranging from about 0.5 SDs above the mean to above 3.00 SDs above the mean, the amount of test information was at least 24 (which yields a standard error of estimate about 0.8). Marginal reliability was equal to or greater than 0.96 within the range described. The reliability between about −0.5 SDs below the mean and above 3 SDs above the mean was .90.

Fig. 3 illustrates the test information function (TIF) represented by a solid line for the 22-item NAQ-R measure. The TIF indicates that the NAQ-R measure yields relatively consistent information, averaging around 24, within a range of approximately 0.5 standard *SD*s from the mean up to over 3 *SD*s above the mean. This range exhibits a marginal reliability of about .96 and an expected standard error of estimate, represented by the dashed line in Fig. 3, of approximately 0.2 for scores within this interval. The marginal reliability for response pattern scores, as provided by IRTPRO, was estimated at .89 for the entire continuum. For the abbreviated versions, the 13-item NAQ-R-SF exhibited a marginal reliability of .82, while the 9-item S-NAQ had a marginal reliability of .79. These marginal reliability values are approximations spanning the entirety of the continuum.

### Is there differential item functioning observed between males and females?

We did not observe sex-related differences in the interpretation of the items. However, in the NAQ-R, Item 3 (“Being ordered to do work below your level of competence”) exhibited minor DIF with a lower discrimination parameter (*a*) in males (0.68) compared to females (1.09), suggesting it is more indicative of WB in females. Notably, we have previously noted that Item 3 should be considered for removal from the NAQ-R item set due to its nonmonotonicity. Similarly, Item 9 (“Intimidating behaviors such as finger-pointing, invasion of personal space, shoving, blocking your way”) displayed minor DIF, with a distinct location parameter (*b*) observed in males (*p* = .041). In the NAQ-R-SF, Item 9 exhibited minor DIF, with a distinct location parameter (*b*) observed in males (*p* = .048). After correcting for multiple testing with Bonferroni adjustment, no significant sex-related DIF was observed in either the classical 22-item NAQ-R or the 13-item NAQ-R-SF developed in this study. No sex-related DIF was found in the S-NAQ.

### Additional comparative and correlational analyses

#### Confirmatory factor analyses of single factor models

All free models for WB fitted well (Table 2). The 22-item NAQ-R model exhibited inferior fit indices compared to both the 9-item S-NAQ and the 13-item NAQ-R-SF models, thus affirming the structural validity of the abbreviated versions. The results underscore that both the 9-item S-NAQ and the 13-item NAQ-R-SF showed approximately similar fit statistics, and better compared to the 22-item NAQ-R (however, see Table 1 for evidence that two items in the S-NAQ were less informative, contrasting with the NAQ-R-SF, which exclusively includes informative items).

**Table 2.** CFA of single-factor models of the NAQ-R, S-NAQ, and the NAQ-R-SF.

| Model | $\chi^2$ (df) | $\chi^2$/df | CFI | SRMR | RMSEA (90% CI) |
| --- | --- | --- | --- | --- | --- |
| 22-item NAQ-R<sup>a</sup> | 1070.45 (205) | 5.22 | .91 | .052 | .070 (.066; .074) |
| 9-item S-NAQ | 95.98 (27) | 3.56 | .98 | .025 | .054 (.043; .066) |
| 13-item NAQ-R-SF<sup>b</sup> | 188.55 (64) | 4.01 | .97 | .030 | .059 (.052; .067) |
| Recommended cut-off point |  | < 5.0 | > .95 | < .08 | < .08 |

*Note.* $N = 867$ . NAQ-R = Negative Acts Questionnaire-Revised. S-NAQ = Short Negative Act Questionnaire. NAQ-R-SF = Negative Acts Questionnaire-Revised-Short Form (developed in this study). <sup>a</sup>Allowed correlation of errors stemmed from two primary sources: proximity of items location (items 3x4) and similarity of content (items 16x21 “unmanageable workload” and “deadline”; items 9x22 “intimidating behavior” and “threats of violence”, and 15x20 items “jokes” and “sarcasm”). <sup>b</sup>Error correlations between items 15x20. $\chi^2$ = Chi square; $p$ of all values is < .001. $\chi^2$ /df = minimum discrepancy divided by its degree of freedom CMIN/df. CFI = confirmatory fit index. SRMR = standardized root mean square residual. RMSEA = root mean square error of approximation. 90%CI = 90% confidence interval.

#### Concurrent validity

Table 3 presents the results of the concurrent validity test, including correlations of the NAQ-R-SF with a single-item measure of WB, the NAQ-R, and the S-NAQ, alongside descriptive statistics. The table indicates good concurrent validity and reliability for the NAQ-R-SF when compared with established and validated measures. Statistical tests were conducted to assess differences in correlation values among the measures (*r* = .53, .52, .57), with the nonsignificant result (*z* = .39, *p* = .349) suggesting a strong level of agreement between these concurrent measures of WB.

**Table 3.**
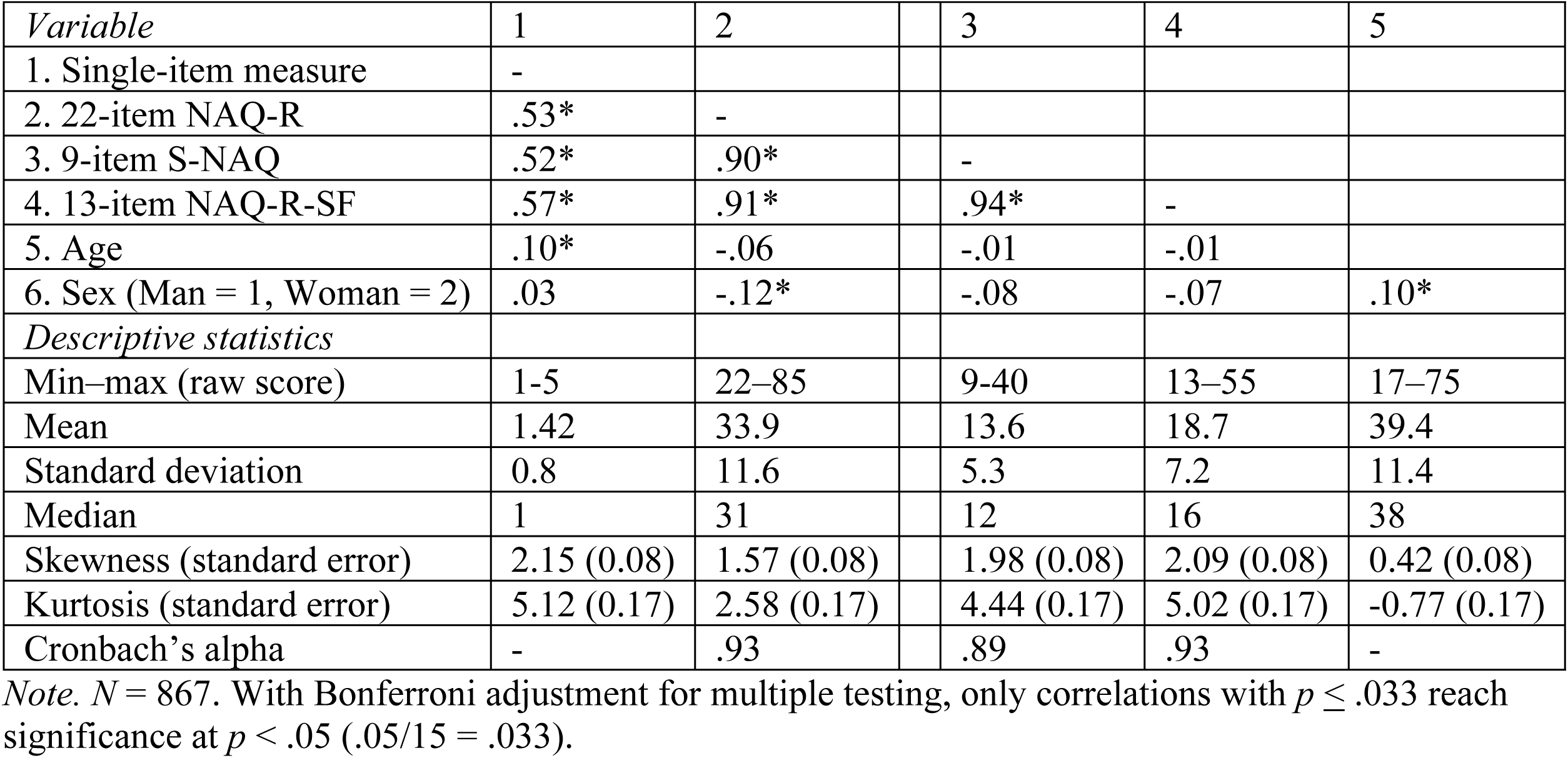
Spearman’s correlation analysis and descriptive statistics for study variables.

#### Divergent validity with individual differences in experiencing workplace bullying

After applying a Bonferroni-adjusted *p*-value for two comparisons (.05/2 = .025), the NAQ-R-SF showed significant negative correlations with both the EQ-5D-3L Index (*r* = −.22) and the EQ-5D-3L VAS (*r* = −.22). For six personality trait comparisons, a Bonferroni-adjusted *p*-value of .008 (.05/6) was used. The NAQ-R-SF demonstrated significant positive correlation with neuroticism (*r* = .27) and significant negative correlations with extraversion (*r* = −.12), agreeableness (*r* = −.13), conscientiousness (*r* = −.14), and honesty-humility (*r* = −.12), but no significant correlation with openness (*r* = −.09). These results support the divergent validity of the NAQ-R-SF.

#### Convergent and divergent validity with variations in workplace functioning

The NAQ-R-SF was significantly positively correlated with interpersonal conflicts at work (*r* = .55) and counterproductive work behavior (CWB) (*r* = .29). After applying a Bonferroni-adjusted *p*-value for three comparisons (.05/3 = .017), it also showed significant negative correlations with task-related work performance (*r* = −.38) and contextual-related work performance (*r* = −.18). These findings provide strong evidence of convergent validity for the NAQ-R-SF (*r* > .50 with interpersonal conflicts at work) and adequate divergent validity (*r* < .50 with other measures of workplace functioning).

## Discussion

The study is groundbreaking in its utilization of several applications of IRT, allowing for a comprehensive psychometric analysis of the 22-item and 9-item NAQ-R measures at both item and scale levels. Additionally, it facilitated the development of a concise 13-item WB measure (NAQ-R-SF), and investigated potential sex-related DIF. Applying classical CTT (CFA, correlations) we validated the NAQ-R-SF across a substantial and well-defined employee sample.

Our study extends the understanding of the NAQ-R through the application of IRT, an approach not widely explored in prior NAQ-R studies. For instance, Caponecchia and Costa [19] primarily utilized IRT for analyzing items in relation to the response format. Similarly, Ma et al. [49] applied IRT but focused on computerized adaptive testing among nurses in a specific cultural context, which makes direct comparisons challenging.

The development of a short version of an instrument requires that the theoretical rationale of the original instrument is well represented in the shortened version. We acknowledge that any short form should adhere to this theoretical rationale; otherwise, it cannot be considered a true short form of the NAQ-R as it would operationalize a different construct. In line with this view, both short versions (S-NAQ and NAQ-R-SF) include items reflecting the three forms of WB (work-related, person-related, and physically intimidating) described by Einarsen et al. [11], though these forms are represented by different items. (See Table 1 for a detailed breakdown of the items included in both the S-NAQ and NAQ-R-SF.) Through IRT analysis, it was revealed that the most informative items within the 9-item S-NAQ capture person-related WB. Notelaers et al. [12] aimed to develop a short measure encompassing different forms of WB. However, in the S-NAQ, only Item 1 (“Someone withholding information which affects your performance”) reflects work-related bullying, which was found not informative in our IRT analysis (see Table 1). Similarly, only Item 8 (“Being shouted at or being the target of spontaneous anger”) reflects physically intimidating bullying, and it was less informative in our IRT analysis. These two items showed lower factor loadings (< .70), consistent with the argument by Notelaers et al. [12] that items reflecting physically intimidating bullying consistently exhibit lower factor loadings, suggesting physical aggression may not constitute WB. As a result of performed IRT analysis, we excluded two of the three items reflecting physically intimidating bullying from the NAQ-R-SF. We acknowledge that physical aggression encompasses various constructs, and its more severe manifestations, as exemplified in item 22 (“Threats of violence or physical abuse or actual abuse”), are governed by distinct laws compared to WB. This particular item has been removed from the two abbreviated versions. However, Item 9 (“Intimidating behaviors such as finger-pointing, invasion of personal space, showing, blocking your way”) exhibited favorable IRT parameters, suggesting its inclusion in our condensed version, NAQ-R-SF. In conclusion, the item selection process for our developed short version of the NAQ-R, denoted NAQ-R-SF, utilized a method, IRT, specifically recommended for scale reduction. Although these two short versions of the NAQ-R were shortened by different methods, NAQ-R-SF maintains a comparable proportion of items to the S-NAQ, encompassing the majority of items related to person-related WB.

Despite employing distinct strategies for abbreviation, both versions serve as concise tools for evaluating WB, encapsulating the three forms of negative behaviors measured by the NAQ-R. However, only 2 out of 13 items (NAQ-R-SF) and 2 out of 9 items (S-NAQ) pertain to the other two forms of WB, thus, the work-related bullying dimension is minimally represented in both short scales. This may raise concerns that these short scales may not be true abbreviations of the NAQ-R, as the proportion of items representing the three forms and their contribution to the total score has changed. However, Notelaers et al. [12] cautioned bullying researchers to be mindful when differentiating between dimensions of WB. While various types of negative social behaviors exist, their findings indicated that this does not imply a clear distinction between different forms of bullying itself. Our analysis shows that both short scales are unidimensional and fit the data well. The possible labeling of these WB forms is misleading, as most items related to person-related WB focus on social isolation at the workplace (e.g., Item 6, “Being ignored or excluded” or Item 12, “Being ignored or facing a hostile reaction when you approach”), which remains highly relevant for employee satisfaction and performance. Notably, in the current study, the NAQ-R-SF showed significant correlations with variables measuring person-related constructs, such as health quality and personality, as well as work-related constructs, such as interpersonal conflicts at work and work performance. This supports the idea that the NAQ-R-SF captures a broad range of WB behaviors.

In our study, after adjusting for multiple testing, we found that sex-related DIF was not significant. This finding aligns with prior research by Sischka et al. [17], suggesting that men and women interpret items related to experienced WB similarly.

In summary, we successfully validated the new NAQ-R-SF. The structural validity of the NAQ-R-SF demonstrated similar fit statistics compared to the S-NAQ and NAQ-R (see Table 2). Like the NAQ-R and S-NAQ, the NAQ-R-SF includes items that reflect all three forms of workplace bullying and are similarly interpreted by men and women. It also exhibited appropriate concurrent, convergent and divergent validity, consistent with theoretical expectations and previous research [3,4,6–8,31], supporting the notion that personality is associated with WB [9,10,18,23,26].

### Limitations and future research

The study’s limitations echo common issues in psychological research, including the treat of Likert-like scale items as approximately continuous, self-report biases and constraints due to the study’s cross-sectional design. Nonetheless, the study’s strengths lie in the focused sample of employed persons, a sizable participant pool, and the utilization of IRT, ensuring a high degree of reliability.

NAQ-R data are based on a five-point Likert-like measure, which we treat as approximately continuous when performing statistical analyses such as CFA. Utilizing Likert-like data consistently supports treating these variables as approximately continuous in both applied and organizational psychology. While technically ordinal, Likert-like scales comprise ordered categories. However, it is worth noting that this approach has faced criticism. Liddell and Kruschke [50] conducted an extensive survey of articles across prominent psychology journals, revealing that all studies examining ordinal data employed a metric model. However, this theoretical discrepancy is, according to Norman [20], irrelevant to the analysis since the computer lacks the capacity to confirm or deny it. Additionally, Robitzsch [51] emphasized the complexity of determining the appropriate modeling strategy for ordinal variables in factor analysis.

This study utilized a convenience sample for efficient data collection, but its generalizability may be restricted due to the non-random sampling approach. Future research should utilize probability sampling techniques to further validate and extend these findings. Despite challenges in designing ideal studies on WB, we condensed the NAQ-R to a 13-item measure using data from Swedish organizations sampled between 2015 and 2019, incorporating both paper-and-pencil and electronic methods, with a predominance of female participants. Similarly, Notelaers et al. [12] shortened the NAQ-R to a 9-item measure, sampling data from Belgian organizations between 2008 and 2016, incorporating both paper-and-pencil and electronic methods, with a predominance of male participants. NAQ-R-SF was constructed using Swedish data, potentially reflecting cultural influence. Despite its favorable IRT parameters, we opted to exclude Item 19. Notably, this item, as highlighted by Notelaers et al. [12], was discussed at international conferences but was not included in the 9-item version (S-NAQ).

Future research should focus on further validating the 13-item NAQ-R-SF by incorporating variables such as sickness absenteeism, presenteeism, recovery, and job satisfaction. Including these variables would offer a more comprehensive understanding of the measure’s effectiveness and applicability across diverse contexts.

## Conclusions

Based on our findings, we conclude that both short measures of WB (9-item S-NAQ, and 13-item NAQ-R-SF) are suitable for both research and practical applications. However, the NAQ-R-SF, introduced in this study, may be preferred due to its exceptional item properties. The NAQ-R-SF proves particularly advantageous for researchers and practitioners aiming to apply it as a continuous assessment tool. Conversely, the S-NAQ may be more useful for researchers applying latent class analysis. IRT and validity evidence support the NAQ-R-SF as a robust tool for measuring WB, aligning with established WB constructs and individual differences.

## Data Availability

The data underlying the results presented in the study are available from Mendeley (doi: 10.17632/mgwtzjww7g.1)

## Acknowledgments

We express our gratitude to Seburan Aliti, Mathilde Faure Lindh, Jennifer Fransson, Magdalena Palander, Carina Ragnestål-Impola, Valentina Tesouri, Davina Tesouza for their invaluable assistance with data sampling; and Björn Persson for performing the Mokken analysis.

